# Predicting the time course of replacements of SARS-CoV-2 variants using relative reproduction numbers

**DOI:** 10.1101/2022.03.30.22273218

**Authors:** Chayada Piantham, Kimihito Ito

## Abstract

The severe acute respiratory syndrome coronavirus 2 (SARS-CoV-2) has continuously evolved since its introduction to the human population in 2019. Natural selection selects variants with higher effective reproduction numbers, increasing the overall transmissibility of the circulating viruses. In order to establish effective control measures for a new variant, it is crucial to know its transmissibility and replacement time course in early phases of the variant replacement. In this paper, we conduct retrospective prediction tests of the variant replacement from Alpha to Delta in England. Our method firstly estimated the relative reproduction number, the ratio of the reproduction number of a variant to that of another, from partial observations up to a given time point. Secondly, the replacement time course after the time point was predicted based on the estimates of relative reproduction number. Thirdly, the estimated relative reproduction number and the predicted time course were evaluated by being compared to those estimated using the entire observations. We found that it is possible to estimate the relative reproduction number of Delta with respect to Alpha when the frequency of Delta was more than or equal to 0.25. Using these relative reproduction numbers, predictions targeting on 1^st^ June 2021, the date when the frequency of Delta reached 0.90, had maximum absolute prediction errors of 0.015 for frequencies of Delta and 0.067 for the average relative reproduction number of circulating viruses with respect to Alpha. These results suggest that our method allows us to predict the time course of variant replacement in future from partial datasets observed in early phases of variant replacement.

## Introduction

Since its first emergence in the human population in 2019, the severe acute respiratory syndrome coronavirus 2 (SARS-CoV-2) has been generating new variants. Natural selection selects new variants that has higher effective reproduction numbers than other circulating variants. As a result, the average transmissibility in the viral population increases over time [1]. As of date, Alpha (B.1.1.7), Beta (B.1.351), Gamma (P.1), Delta (B.1.617.2), and Omicron (B.1.1.529) have been assigned as variants of concern (VOCs) because of their increased transmissibility compared to previous variants [2].

It is important to know how transmissible the new variants are compared to previously circulating variants because the average reproduction number of the circulating virus changes when new variants become dominant. Several studies have analyzed the reproduction numbers of new variants that have replaced old ones. Volz et al. estimated the effective reproduction number of Alpha in England to be 1.5–2.0 times higher than that of non-VOCs using a logistic growth model for variant frequencies [3]. Leung et al. estimated the basic reproduction number of Alpha to be 1.75 times higher than that of previously circulating variants in England using a renewal-equation-based model [4]. Ito et al. estimated the effective reproduction number of Delta to be 1.35 times higher than that of Alpha from variant frequencies observed in Japan by using an approximated version of the renewal-equation-based model [5]. Using the same method, Ito et al. estimated the effective reproduction number of Omicron to be 3.15 times higher than that of Delta in Denmark [6], and Nishiura et al. estimated the effective reproduction of Omicron to be 4.2 times higher than that of Delta in South Africa [7].

In order to prepare control measures against new variants, it is crucial to predict the time course of the variant replacements in advance. The prediction of variant selection has been widely studied in seasonal influenza viruses [8]. Łuksza and Lässig developed a fitness model using mutations on epitopes and non-epitopes to predict selected variants [9]. Huddleston et al. predicted the future frequency of variants using its current frequency, the antigenic novelty of epitopes, and the mutational load in non-epitopes [10]. Piantham and Ito modeled the fixation probability of variants using variant frequency and statistics on patient ages [11]. In the case of seasonal influenza, the main driving force of natural selection was the population immunity acquired from previous infections. In contrast, most of the human population are considered naïve to SARS-CoV-2 infections, and a new method to predict the time course of variant replacement of SARS-CoV-2 needs to be developed.

The transmissibility of an infectious agent can be measured by its reproduction number. The effective reproduction number at time *t* (*R*_*t*_) is defined as the average number of people someone infected at time *t* could expected to produce if conditions should remain unchanged [12]. When more than one variant of the infectious agent is circulating, the relative reproduction number can be used to measure the relative transmissibility of a variant compared to a baseline variant [4,13]. However, the method requires the number of new cases in addition to the frequencies of variants, and it is not applicable for predicting future time course of variant replacement. Using approximations, Ito et al. proposed a method to determine the relative reproduction number without knowing the number of new cases [5]. This method allows us to predict the future time course of variant replacements.

Nucleotide sequences of SARS-CoV-2 variants collected worldwide have been submitted to and accumulated on the GISAID database [14]. It is known that different geographical locations have different distributions of variants [15]. As of 28^th^ February 2022, a total of 8,753,735 sequences have been registered on the database worldwide. Of these, 1,786,080 (20%) were submitted from England, which has their population account for 0.71% of the world population. These numbers indicate that England is one of the locations having highest sequencing capacity. In England, the Alpha–Delta replacement was observed from March 2021 to June 2021. The sequence information during the Alpha–Delta replacement in England is one of the best datasets to evaluate the predictability of variant replacement in SARS-CoV-2.

In this study, we conduct retrospective prediction tests using the nucleotide sequences collected in England during the Alpha–Delta replacement. For each given time point, we use partial sequence data observed only up to that time point to estimate the relative reproduction number of Delta with respect to (w.r.t.) Alpha. The estimated relative reproduction number is then used to predict the future time course of variant replacement. The estimated relative reproduction numbers and the predicted time courses are evaluated by being compared to those estimated using the entire dataset.

## Materials and Methods

### Nucleotide sequences

Nucleotide sequences of SARS-CoV-2 viruses collected from England during 1^st^ January 2021 to 31^st^ July 2021 were downloaded from the GISAID database on 16^th^ November 2021 [14]. Of these, 411,123 sequences had complete information about date of sample collection in the metadata. The PANGO lineage names [16] of these sequences were collected from metadata and recorded with their collection dates (Supplementary Table S1). Sequences that are labeled as “B.1.1.7” or sublineage names starting with “Q.” were classified as the Alpha variant. Sequences that are labeled as “B.1.617.2” or sublineage names starting with “AY.” were classified as the Delta variant. There were 11,773 sequences of lineages other than Alpha and Delta, and these were ignored in subsequent analyses. A total of 399,350 sequences of Alpha (192,250) and Delta (207,100) were used for counting the daily numbers of sequences belonging to Alpha and Delta (Supplementary Fig. S1).

### Model of Advantageous Selection

We estimated the relative reproduction number of a variant w.r.t. a baseline variant using an approximated version of the renewal-equation-based model [5], which is based on Fraser’s time-since-infection model [12]. Briefly, suppose the viral population consisted of only variant *X* at time *t*_0_. Variant *Y* was introduced into the population at calendar time *t*_*Y*_ > *t*_0_ with an initial frequency of *q*_*Y*_ (*t*_*Y*_). Note that *q*_*Y*_(*t*) = 0 for *t*_0_ ≤ *t* < *t*_*Y*_. The date of variant introduction, *t*_*Y*_, was set to the first date when more than one sequence of that variant was counted. We assume the effective reproduction number of variant *Y* was *k* times higher than that of variant *X*. Let *f*(*τ*) be the probability mass function of generation time *τ*. We assume *f*(*τ*) follows the gamma distribution with a shape parameter of 3.42 and a scale parameter of 1.36 [17]. We discretize *f*(*τ*) to 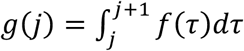 for 1 ≤ *j* ≤ 20. We truncate the generation time distributions at *τ* = 1 and *τ* = 20 by adding 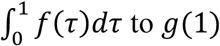 to *g*(1) and 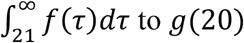 to *g*(20) so that 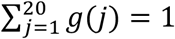. Let *I*(*t*) be the total number of new infections by either *X* or *Y* at calendar time *t*, the effective reproduction numbers of variant *X* and *Y* can be calculated as

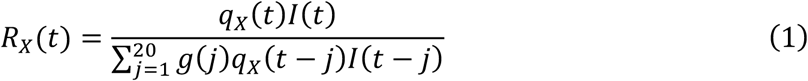

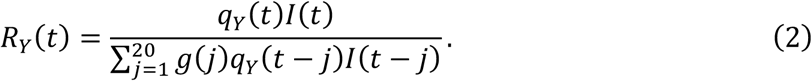

Since the effective reproduction number of variant *Y* is *k* times higher than that of variant *X*, the effective reproduction number of variant *Y* at time *t* is given by

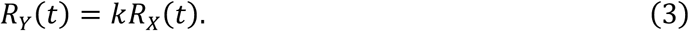

Assuming that the viral population at time *t* comprises of only variants *X* and *Y*, the frequency of variant *Y* at calendar time *t, q*_*Y*_(*t*), can be calculated as

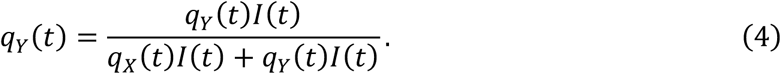

We assume that the numbers of new infections do not vary greatly for 20 days, i.e.

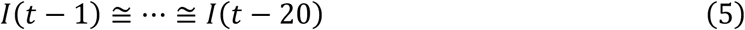

for *t* > *t*_0_. Using this approximation with Equations (1), (2), and (3), we can rewrite Equation (4) using *q*_*Y*_(*t* − *j*) for 1 ≤ *j* ≤ 20 as

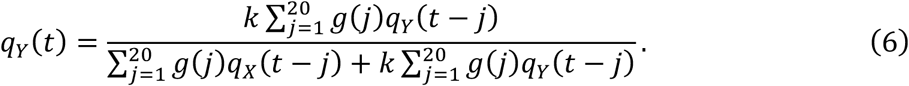

The average relative reproduction number of circulating viruses at time *t* w.r.t. variant *X* is given by

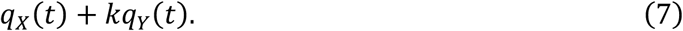

### Parameter estimation from the number of sequences

Let *N*_*X*_(*t*) and *N*_*Y*_(*t*) be the number of sequences of variant *X* and *Y* observed at calendar time *t*, respectively. Suppose that variant *Y* is sampled and sequenced following a beta-binomial distribution having distribution parameters of *α* = *q*_*Y*_(*t*)*M* and *β* = (1 − *q*_*Y*_(*t*))*M*. Note that this beta-binomial distribution has a mean of (*N*_*X*_(*t*) + *N*_*Y*_(*t*))*q*_*Y*_(*t*) and a variance of 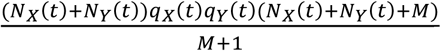. The beta-binomial distribution becomes the binomial distribution when *M* = ∞. The following equation gives the likelihood function of parameters *k, q*_*Y*_(*t*_*Y*_), and *M* for observing *N*_*X*_(*t*) and *N*_*Y*_(*t*) sequences of variant *X* and *Y* at calendar time *t*:

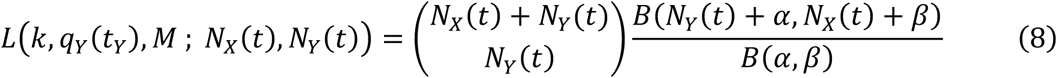

where *α* = *q*_*Y*_(*t*)*M, β* = (1 − *q*_*Y*_(*t*))*M*, and 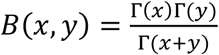.

The likelihood for observing *N*_*Y*_(*t*) sequences of variant *Y* during the period on calendar times *t*_1_, …, *t*_*n*_ is given by the product of the above formula for 1 ≤ *t* ≤ *n*. The estimates of *k, q*_*Y*_(*t*_*Y*_), and *M* were obtained by maximizing the likelihood function using observed numbers of sequences in England from *t* = *t*_*Y*_ until the latest *t* in which *q*_*Y*_(*t*) < 1. The 95% confidence intervals (95% CI) of parameters were determined using the profile likelihood method [18]. From the maximum likelihood estimates of *k* and *q*_*Y*_(*t*), the average relative reproduction number of circulating viruses w.r.t. Alpha at time *t* was estimated from Equation (7).

### Prediction of variant frequency and average relative reproduction number

Frequencies of Delta and average relative reproduction numbers of circulating viruses w.r.t. Alpha in future were predicted using the maximum likelihood estimates of parameters calculated from early observations. First, we estimated *k, q*_*Y*_(*t*_*Y*_), and *M* using all observations of *N*_*X*_(*t*) and *N*_*Y*_(*t*) for *t*_*Y*_ ≤ *t* ≤ *t*_*n*_ where *t*_*n*_ is the latest calendar time *t* such that *q*_*Y*_(*t*) < 1, by maximizing likelihood of products of Equation (8). Using the maximum likelihood estimates and their 95% CIs, we determined the calendar times *t* when the estimated frequency *q*_*Y*_(*t*) exceeded 0.05, 0.10, 0.15, 0.20, 0.25, 0.30, 0.35, 0.40, 0.45, 0.50, 0.55, 0.60, 0.65, 0.70, 0.75, 0.80, 0.85, 0.90, and 0.95. For each date *t* determined above, we calculated the maximum likelihood estimates of *k, q*_*Y*_(*t*_*Y*_), and *M* using observations no later than *t*. Frequencies of Delta and average relative reproduction numbers of circulating viruses w.r.t. Alpha in future were predicted by substituting *k* and *q*_*Y*_(*t*_*Y*_) in Equations (6) and (7), respectively.

## Results

### Estimation of relative reproduction number from entire observations

Parameters of the model were estimated using the entire observations from 18^th^ March to 4^th^ July 2021 in England (Table 1). The relative reproduction number (*k*) of Delta w.r.t. Alpha was estimated to be 1.67 (95% CI: 1.64, 1.67) with a beta-binomial distribution parameter (*M*) of 286.82 (95% CI: 201.83, 403.22).

**Table 1.** Parameters estimated using the entire observations during the Alpha–Delta replacement.

| $k$ (95% CI) | $q_Y(t_Y)$ (95% CI) | $M$ (95% CI) | Log Likelihood |
| --- | --- | --- | --- |
| 1.67 (1.64, 1.67) | 0.0005 (0.0004, 0.0006) | 286.82 (201.83, 403.22) | −430.97 |

Figure 1a shows the observed and estimated frequencies of Delta during the Alpha–Delta replacement in England. The blue curve and black dots around the blue curve represent the maximum likelihood estimates and 95% CI of frequencies of Delta. The gray area represents 95% equal-tailed intervals of the beta distribution with the parameters *q*_*Y*_(*t*) and *M* in the 95% CI. Figure 1b shows the maximum likelihood estimations and 95% CI of the average relative reproduction number of circulating viruses w.r.t. Alpha during the same period. Dashed vertical lines in both panels indicate the dates when frequencies of Delta exceeded each 0.05 increment from 0.05 to 0.95 (Table 2). It took 48 days for Delta to reach from frequencies of 0.05 to 0.95.

**Table 2.**
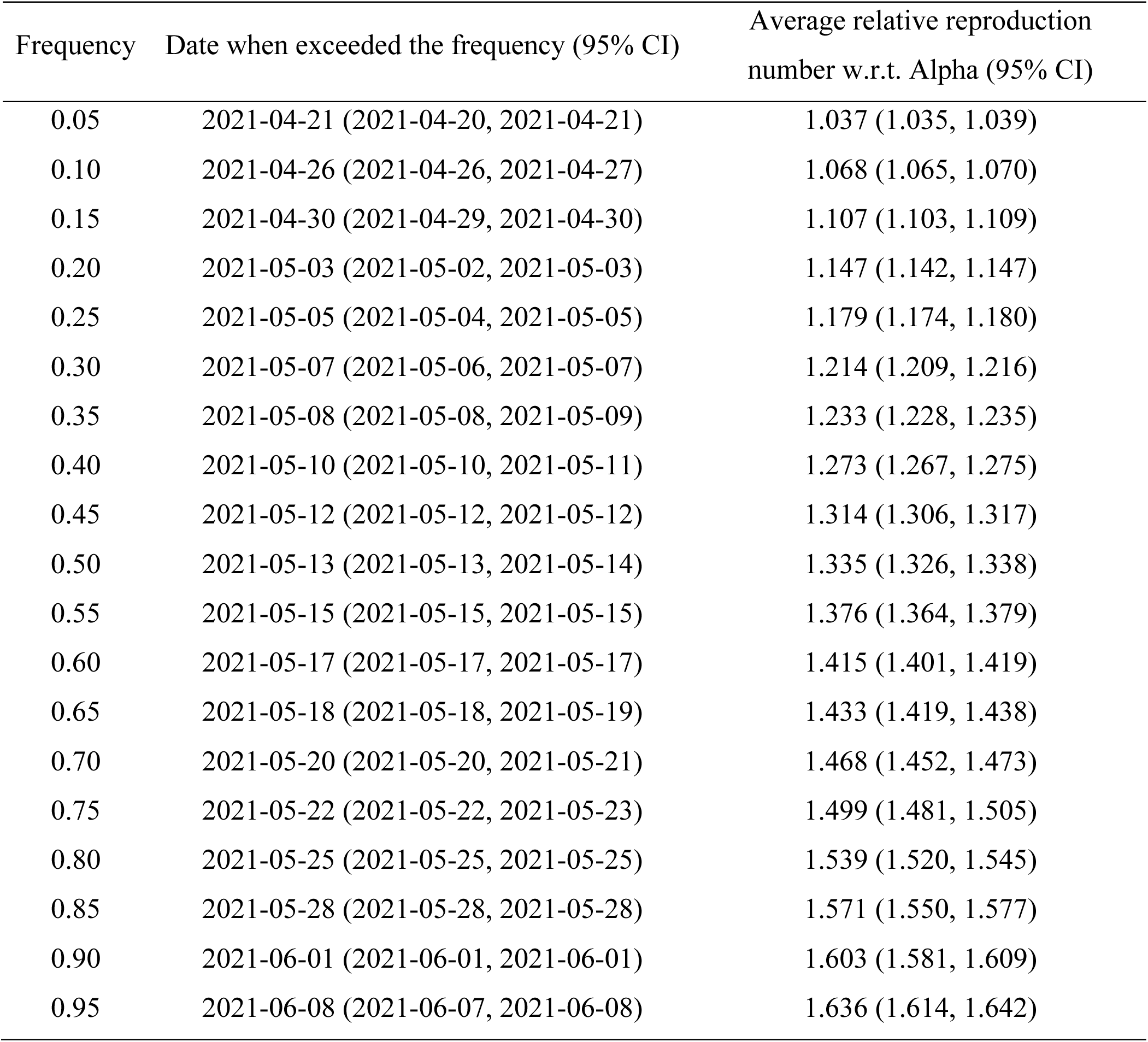
Maximum likelihood estimations for dates when Delta exceeded certain frequencies and the average relative reproduction numbers w.r.t. Alpha on those dates.

| Frequency | Date when exceeded the frequency (95% CI) | Average relative reproduction number w.r.t. Alpha (95% CI) |
| --- | --- | --- |
| 0.05 | 2021-04-21 (2021-04-20, 2021-04-21) | 1.037 (1.035, 1.039) |
| 0.10 | 2021-04-26 (2021-04-26, 2021-04-27) | 1.068 (1.065, 1.070) |
| 0.15 | 2021-04-30 (2021-04-29, 2021-04-30) | 1.107 (1.103, 1.109) |
| 0.20 | 2021-05-03 (2021-05-02, 2021-05-03) | 1.147 (1.142, 1.147) |
| 0.25 | 2021-05-05 (2021-05-04, 2021-05-05) | 1.179 (1.174, 1.180) |
| 0.30 | 2021-05-07 (2021-05-06, 2021-05-07) | 1.214 (1.209, 1.216) |
| 0.35 | 2021-05-08 (2021-05-08, 2021-05-09) | 1.233 (1.228, 1.235) |
| 0.40 | 2021-05-10 (2021-05-10, 2021-05-11) | 1.273 (1.267, 1.275) |
| 0.45 | 2021-05-12 (2021-05-12, 2021-05-12) | 1.314 (1.306, 1.317) |
| 0.50 | 2021-05-13 (2021-05-13, 2021-05-14) | 1.335 (1.326, 1.338) |
| 0.55 | 2021-05-15 (2021-05-15, 2021-05-15) | 1.376 (1.364, 1.379) |
| 0.60 | 2021-05-17 (2021-05-17, 2021-05-17) | 1.415 (1.401, 1.419) |
| 0.65 | 2021-05-18 (2021-05-18, 2021-05-19) | 1.433 (1.419, 1.438) |
| 0.70 | 2021-05-20 (2021-05-20, 2021-05-21) | 1.468 (1.452, 1.473) |
| 0.75 | 2021-05-22 (2021-05-22, 2021-05-23) | 1.499 (1.481, 1.505) |
| 0.80 | 2021-05-25 (2021-05-25, 2021-05-25) | 1.539 (1.520, 1.545) |
| 0.85 | 2021-05-28 (2021-05-28, 2021-05-28) | 1.571 (1.550, 1.577) |
| 0.90 | 2021-06-01 (2021-06-01, 2021-06-01) | 1.603 (1.581, 1.609) |
| 0.95 | 2021-06-08 (2021-06-07, 2021-06-08) | 1.636 (1.614, 1.642) |

**Figure 1.**
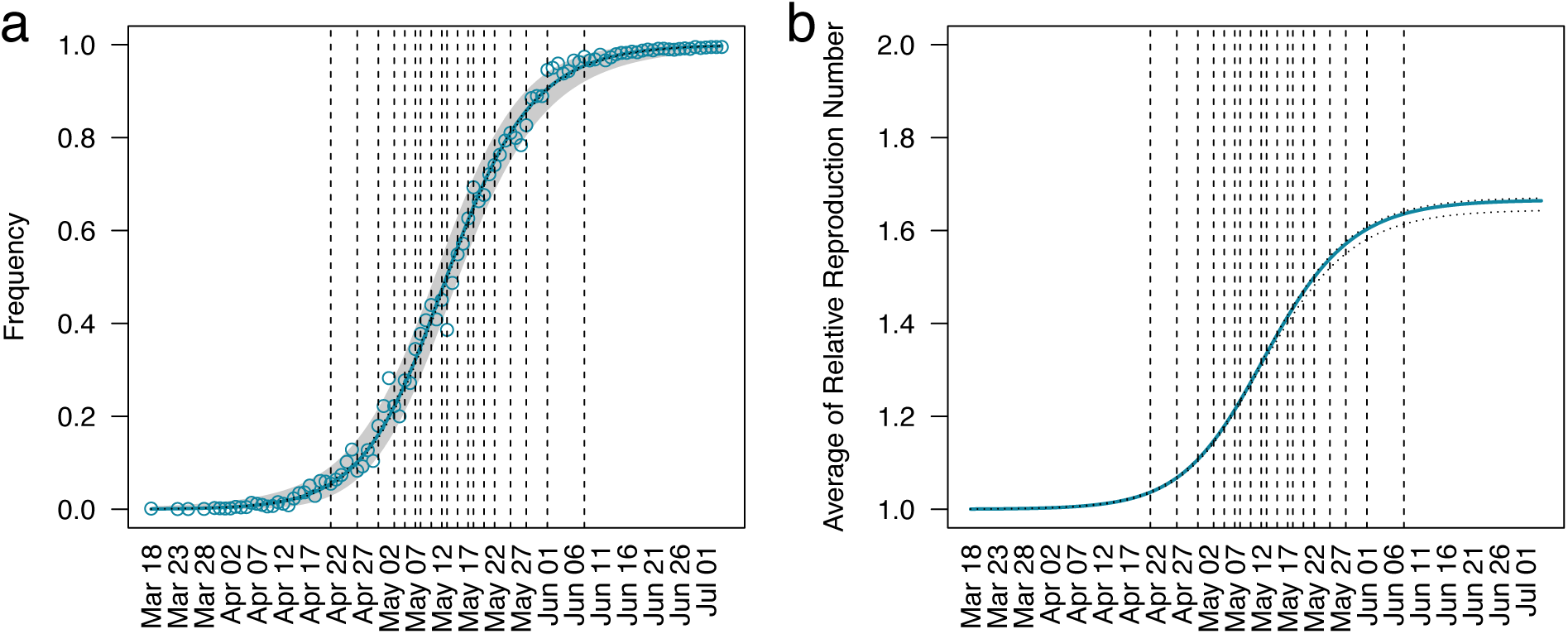
Estimated frequencies of the Delta variant and average relative reproduction number of circulating viruses w.r.t. Alpha during 18^th^ March to 4^th^ July 2021 using entire observations. In panel **a**, circles represent variant frequencies observed in the GISAID database. The blue curve represents the maximum likelihood estimates of frequencies of Delta. Black dotted curves around the blue curve represent 95% confidence intervals of the estimated frequencies of Delta. Gray area represents the 95% equal-tailed interval of beta distribution for the 95% confidence intervals of parameters of the estimated beta-binomial distribution. In panel **b**, the blue curve and black dotted curves represents the maximum likelihood estimates and 95% confidence intervals of the average relative reproduction number of circulating viruses with respect to Alpha. Vertical dashed lines in both panels indicate the dates when the estimated frequency reached 0.05, 0.10, 0.15, 0.20, 0.25, 0.30, 0.35, 0.40, 0.45, 0.50, 0.55, 0.60, 0.65, 0.70, 0.75, 0.80, 0.85, 0.90, and 0.95.

### Estimation of relative reproduction number from partial observations

Table 3 shows the parameters of our model estimated using the partial data collected no later than each maximum likelihood date in Table 2. The maximum likelihood estimate of *k* using observations of the entire period in the Alpha–Delta replacement was 1.67 (Table 1). We call this estimate the ‘final estimate’. The final estimate was within 95% CIs of estimations in six (0.15, 0.25, 0.30, 0.35, 0.40, and 0.45) out of nine estimations using the partial observations before Delta reached frequencies of 0.50. Three early estimations made at frequencies of 0.05, 0.10, and 0.20 failed to cover the final estimate of *k* in their 95% CIs. The five estimations made at frequencies greater than or equal to 0.25 covered the final estimate of *k* in their 95% CIs. This implied that it was possible to accurately estimate the relative reproduction number of Delta w.r.t. Alpha when frequencies of Delta became 0.25 or later. It took 34 days for Delta to reach a frequency of 0.95 from when it was 0.25 (Table 2). Therefore, we would be able to estimate the relative reproduction number of Delta w.r.t. Alpha more than one month before its fixation. The estimated values of *M* also became steady at around 300.00 at frequencies greater than or equal to 0.25 (Table 3).

**Table 3.** Parameters estimated using partial observations.

| Observed<br>frequency | $k$ | $q_0$ | $M$ | Log Likelihood |
| --- | --- | --- | --- | --- |
| 0.05 | 1.78 (1.69, 1.85) | 0.0003 (0.0002, 0.0004) | 611.41 (268.84, 2000.00<sup>†</sup>) | −82.09 |
| 0.10 | 1.76 (1.68, 1.80) | 0.0003 (0.0002, 0.0004) | 559.33 (264.55, 1476.99) | −102.85 |
| 0.15 | 1.69 (1.62, 1.71) | 0.0004 (0.0003, 0.0006) | 395.63 (202.88, 842.16) | −123.12 |
| 0.20 | 1.72 (1.69, 1.73) | 0.0003 (0.0003, 0.0004) | 359.75 (189.17, 725.97) | −137.61 |
| 0.25 | 1.67 (1.62, 1.71) | 0.0005 (0.0003, 0.0007) | 283.92 (160.53, 603.44) | −148.98 |
| 0.30 | 1.68 (1.66, 1.70) | 0.0004 (0.0003, 0.0005) | 313.07 (171.38, 579.86) | −157.87 |
| 0.35 | 1.67 (1.63, 1.74) | 0.0004 (0.0003, 0.0006) | 311.65 (175.27, 602.30) | −161.95 |
| 0.40 | 1.68 (1.64, 1.70) | 0.0004 (0.0004, 0.0006) | 335.32 (189.19, 637.92) | −170.37 |
| 0.45 | 1.67 (1.65, 1.72) | 0.0004 (0.0003, 0.0005) | 319.57 (181.41, 575.63) | −180.60 |
| 0.50 | 1.64 (1.60, 1.69) | 0.0006 (0.0004, 0.0008) | 230.02 (137.25, 411.11) | −190.09 |
| 0.55 | 1.62 (1.59, 1.65) | 0.0007 (0.0005, 0.0008) | 218.52 (133.36, 439.22) | −199.09 |
| 0.60 | 1.64 (1.63, 1.68) | 0.0005 (0.0004, 0.0006) | 254.02 (150.72, 429.33) | −207.23 |
| 0.65 | 1.65 (1.64, 1.69) | 0.0005 (0.0004, 0.0006) | 245.72 (148.15, 413.85) | −213.00 |
| 0.70 | 1.65 (1.63, 1.67) | 0.0005 (0.0004, 0.0006) | 256.17 (154.91, 426.78) | −222.19 |
| 0.75 | 1.64 (1.63, 1.66) | 0.0005 (0.0005, 0.0007) | 264.58 (162.71, 451.18) | −230.71 |
| 0.80 | 1.65 (1.64, 1.68) | 0.0005 (0.0004, 0.0006) | 291.69 (177.99, 484.83) | −244.13 |
| 0.85 | 1.60 (1.59, 1.65) | 0.0008 (0.0005, 0.0009) | 202.38 (119.43, 612.16) | −264.29 |
| 0.90 | 1.65 (1.63, 1.67) | 0.0005 (0.0004, 0.0006) | 236.54 (155.20, 372.12) | −284.01 |
| 0.95 | 1.66 (1.65, 1.68) | 0.0005 (0.0004, 0.0006) | 201.41 (138.15, 306.96) | −328.33 |

<sup>†</sup>The upper bound of $M$ in the maximum likelihood estimation was set to 2,000.

### Prediction of variant frequency and average relative reproduction number

We conducted retrospective prediction tests on the future frequency of Delta and the average relative reproduction number of circulating viruses w.r.t. Alpha using model parameters in Table 3, which were estimated from partial observations. Figure 2 shows predicted trajectories of the Alpha–Delta replacement using partial observations up to different time points in Table 2. The maximum likelihood predictions made at frequencies of 0.05, 0.10, and 0.20 overestimated the future frequencies of Delta (Figures 2a–b, 2d), while predictions made at frequencies greater than or equal to 0.25 fitted well with future observations (Figures 2e–i).

**Figure 2.**
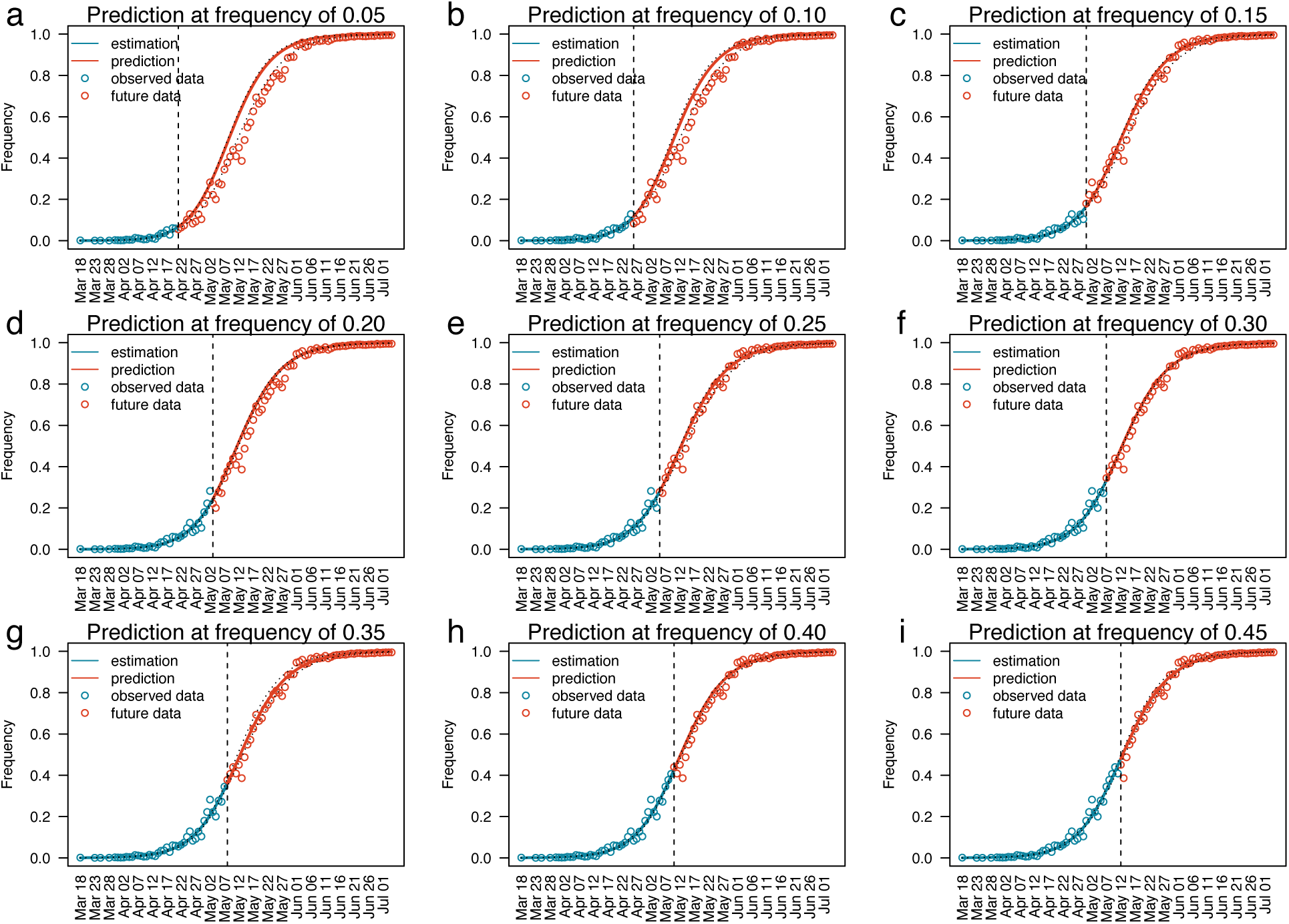
Prediction of future frequencies of the Delta variant using partial observations. Panels **a, b, c, d, e, f, g, h**, and **i** represent predictions estimated using observations until 21^st^ April, 26^th^ April, 30^th^ April, 3^rd^ May, 5^th^ May, 7^th^ May, 8^th^ May, 10^th^ May, and 12^th^ May, 2021, respectively. Blue circles represent observed frequencies used for predictions. Red circles represent future observations that were not used for predictions. The blue curve in each panel represents the maximum likelihood estimates of frequencies, and the red curve represents the frequencies predicted using the estimated parameter. Black dotted curves represent 95% confidence intervals of the estimated frequencies of Delta. The vertical dashed line in each panel represents the date of the last observations used for prediction.

According to the final estimate using the entire observations, Delta exceeded frequencies of 0.50, 0.70, and 0.90 on 13^th^ May, 20^th^ May, and 1^st^ June 2021, respectively (Table 2). We evaluated the accuracy of predictions by analyzing predictions targeted on these dates (Figure 3). As the relative reproduction numbers were overestimated when predictions were made before frequencies of Delta reached 0.25, the frequencies of Delta on the target dates were also overestimated in these predictions (Figure 3a–c). In contrast, predictions made when frequencies of Delta were greater than or equal to 0.25 were close to the final estimate of frequency using entire observations. When frequencies of Delta were below 0.25, the predictions targeted on 13^th^ May, 20^th^ May, and 1^st^ June 2021 had median errors of 0.074, 0.069, and 0.036 with maximum absolute errors of 0.143, 0.121, and 0.055, respectively (Table 4). When frequencies of Delta were greater than or equal to 0.25, the predictions targeted on 13^th^ May, 20^th^ May, and 1^st^ June 2021 had median errors of 0.018, 0.011, and 0.000 with maximum absolute errors of 0.023, 0.023, and 0.015, respectively (Table 5). Predictions made when frequencies of Delta were greater than or equal to 0.25 have significantly smaller prediction errors than those made when frequencies of Delta were less than 0.25 (*p*-values of 0.016, 0.003, and 0.001 using two-sided Wilcoxon rank sum test for predictions targeted on 13^th^ May, 20^th^ May, and 1^st^ June 2021, respectively).

**Figure 3.**
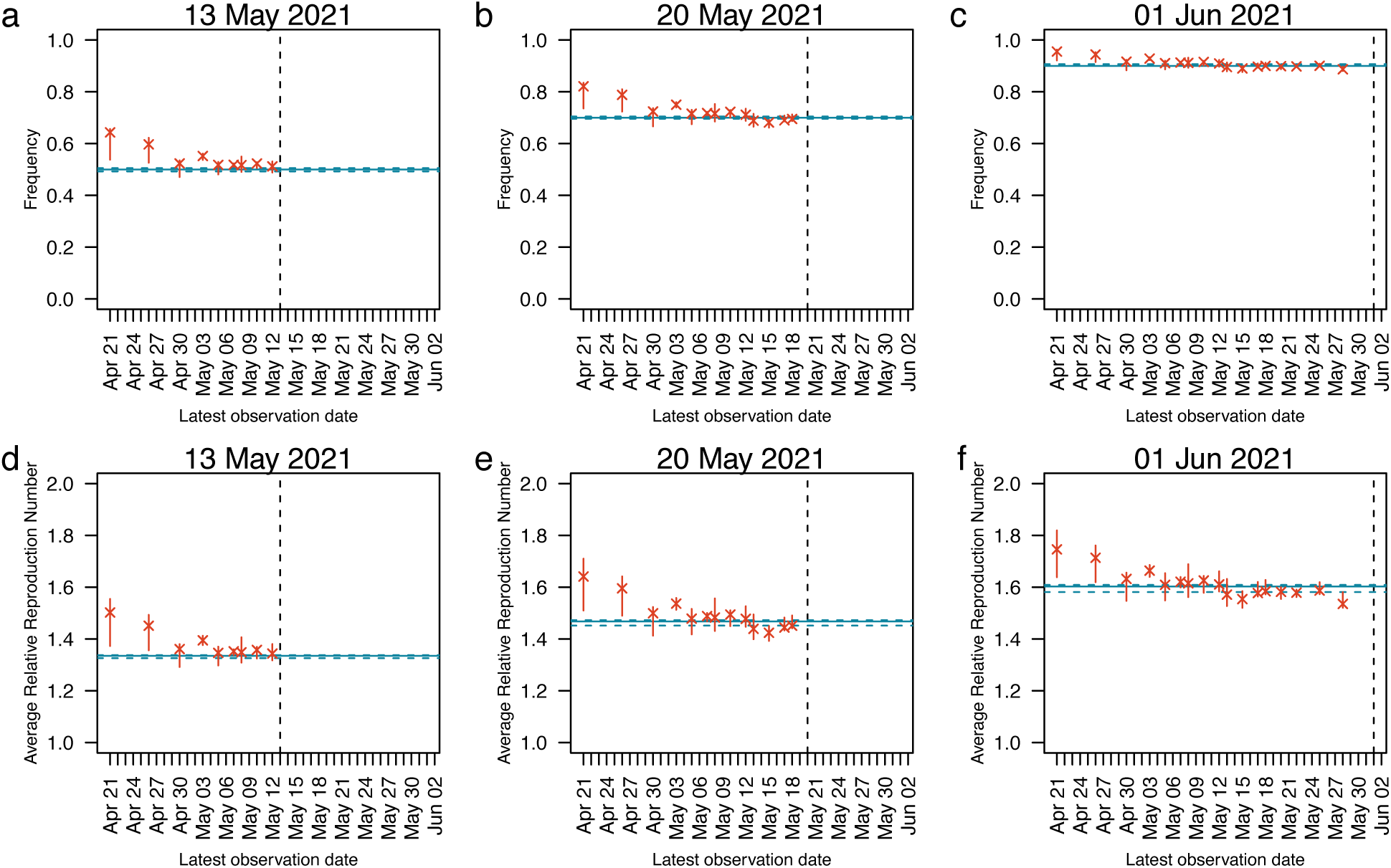
Predictions of population average of relative reproduction number with respect to Alpha. In each panel, x-axis represents dates until which observations were used in the prediction. Y-axis represents the predicted population average of reproduction number for the date marked by vertical dashed lines (13^th^ May in **a**, 20^th^ May in **b**, and 1^st^ June in **c**). Cross marks represent predicted population average of relative reproduction number with respect to Alpha and vertical bars represent their 95% confidence intervals. The blue horizontal solid lines represent the maximum likelihood estimates using the entire observations, we call the final estimates. The horizontal dashed blue lines represent 95% confidence intervals of the final estimates..

**Table 4.**
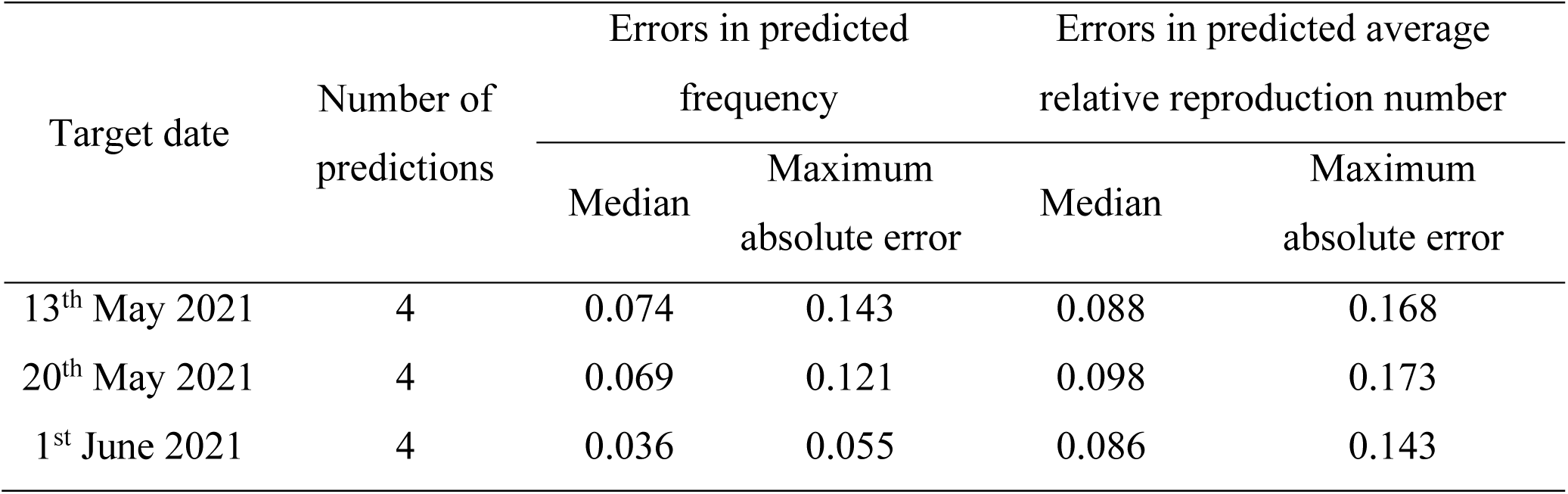
Errors of predictions made at frequencies less than 0.25.

**Table 5.** Errors of predictions made at frequencies greater than or equal to 0.25.

| Target date | Number of predictions | Errors in predicted frequency |  | Errors in predicted average relative reproduction number |  |
| --- | --- | --- | --- | --- | --- |
|  |  | Median | Maximum absolute error | Median | Maximum absolute error |
| 13<sup>th</sup> May 2021 | 5 | 0.018 | 0.023 | 0.014 | 0.023 |
| 20<sup>th</sup> May 2021 | 9 | 0.011 | 0.023 | 0.010 | 0.044 |
| 1<sup>st</sup> June 2021 | 13 | 0.000 | 0.015 | −0.019 | 0.067 |

From the final estimate using entire observations, the average relative reproduction numbers of circulating viruses w.r.t. Alpha on 13^th^ May, 20^th^ May, and 1^st^ June 2021 were estimated to be 1.34, 1.47, and 1.60, respectively (Table 2). In the same way as the frequency of Delta, the average relative reproduction numbers of circulating viruses w.r.t. Alpha on the target dates were overestimated when predictions were made before frequencies of Delta reached 0.25 (Figure 3d– f). When the frequencies of Delta were below 0.25, predictions targeted on 13^th^ May, 20^th^ May, and 1^st^ June 2021 had median prediction errors of 0.088, 0.098, and 0.086 and maximum absolute errors of 0.168, 0.173, and 0.143, respectively (Table 4). When frequencies of Delta were greater than or equal to 0.25, predictions targeted on 13^th^ May, 20^th^ May, and 1^st^ June 2021 had median errors of 0.014, 0.010, and –0.019 with maximum absolute errors of 0.023, 0.044, and 0.067, respectively (Table 5). Predictions made when frequencies of Delta were greater than or equal to 0.25 have significantly smaller prediction errors than those made when frequencies of Delta were less than 0.25 (*p*-values of 0.016, 0.003, and 0.001 using two-sided Wilcoxon rank sum test for predictions targeted on 13^th^ May, 20^th^ May, and 1^st^ June 2021, respectively).

## Discussion

We analyzed the replacement from the Alpha variant to the Delta variant in England using nucleotide sequences on the GISAID database collected between 1^st^ January 2021 to 31^st^ July 2021. The estimated relative reproduction number, *k*, of Delta w.r.t. Alpha was 1.67 (95% CI: 1.64–1.67) with a beta-binomial distribution parameter (*M*) of 286.82 (95% CI: 201.83–403.22) (Table 1). The relative reproduction number was accurately estimated from early observations once frequencies of Delta reached 0.25 (Table 3). Using these relative reproduction numbers, predictions targeting on 1^st^ June 2021, the date when the frequency of Delta reached 0.90, had maximum absolute prediction errors of 0.015 for frequencies of Delta and 0.067 for the average relative reproduction number of circulating viruses with respect to Alpha (Table 5).

Several studies have estimated the relative reproduction of Delta w.r.t. Alpha. Ito et al. estimated the relative reproduction number of Delta w.r.t. Alpha in Japan to be 1.35 [5]. Hansen estimated the relative reproduction number of Delta w.r.t. Alpha in Denmark to be 2.17 [19]. In this study, the relative reproduction number of Delta w.r.t. Alpha was estimated to be 1.67 (95% CI: 1.64– 1.67) (Table 1). Figgins and Bedford found that the relative reproduction number of Delta and Alpha w.r.t. non-VOC variants in the United States were different depending on the states [20]. The differences in relative reproduction numbers of Delta w.r.t. Alpha among countries or states may be attributed to the differences in the vaccine usage or the ethnicity of the target populations.

Our model assumes that the sequences on GISAID database was sampled following a beta-binomial distribution. We could use the binomial distribution in the model instead of the beta-binomial distribution. The model using beta-binomial distribution resulted in lower Akaike information criterions (AIC) compared to the model using only binomial distribution (Supplementary Table S2). This means that the observed variance was larger than the variance of the binomial distribution. The additional variance to the binomial distribution might be attributed to the difference between variant frequencies among subpopulations. For example, different regions may show different progresses in the variant replacement. The same may be true for different age groups. The lower AIC in the beta-binomial distribution model may mean that the target population was not well-mixed.

When a new variant having higher effective reproduction number than the circulating variants emerges, the new variant will be selected by the natural selection. As a result, the average relative reproduction number of the circulating viruses w.r.t. old variants increases. Our retrospective prediction tests show that the average relative reproduction number of circulating viruses w.r.t. Alpha were accurately predicted when observed frequencies of Delta exceeded 0.25. However, some predictions made at higher frequencies (0.55, 0.70, 0.75, and 0.85) were slightly lower than the final estimation using entire observations (Figure 3e–f). One possible reason for these is the relaxation of lockdown restrictions from 17^th^ May 2021. Assuming that variant replacement occurs earlier and sampling is more dense in the city area than countryside, the relaxation of lockdown restrictions mixed viral population in both areas, making the observations of Alpha in city area more than expected.

Prediction tests conducted in this study did not consider delay in sequence submissions. To simulate real-time predictions with submission delays, we conducted the same analysis using observations that were submitted no later than the day of prediction. The results were not as good as the analysis which assumed that there was no submission delay (Supplementary Fig. S2). Even if predictions were made when frequencies of Delta were greater than or equal to 0.25, the frequencies of Delta were overestimated with maximum absolute errors of 0.084, 0.080, and 0.041 for the predictions targeted on 13^th^ May, 20^th^ May, and 1^st^ June 2021, respectively (Supplementary Table S3). The maximum absolute errors of average relative reproduction number of circulating viruses w.r.t. Alpha targeting the same dates were 0.101, 0.113, and 0.099, respectively (Supplementary Table S3). The possible reason for these overestimations is the difference in submission delay between Delta and Alpha sequences. The submission delay of nucleotide sequences belonging to Alpha had a mean of 11.05 days and a median of 10 days during the period from the introduction of Delta (18^th^ March 2021) to the date when Delta reached a frequency of 0.50 (13^th^ May 2021). The submission delay of Delta during the same period had a mean of 10.44 days and a median of 9 days. This means that sequences of Delta were submitted to the GISAID database more quickly than those of Alpha during the early stages of the Alpha–Delta replacement. Sequencing prioritizing a new variant may have led to the overestimations, and is the limitation of our real-time prediction using sequence databases. An explicit inclusion of submission delays for each variant in the model may solve this problem for real-time predictions. Use of data sources without variant prioritization, such as results of PCR tests, can also solve this problem.

Our model assumes that there was no difference between the generation times of both variants with a mean value of 4.64 days [17]. However, Hart et al. estimated the generation time of Delta (4.7 days) to be shorter than that of Alpha (5.5 days) [21]. To allow differences between generation times of variants, it is necessary to extend the model to also estimate the relative generation times of the variant w.r.t. that of the baseline variant [22].

## Data Availability

All data produced in the present work are contained in the manuscript

## Acknowledgement

We gratefully acknowledge the laboratories responsible for obtaining the specimens and the laboratories where genetic sequence data were generated and shared via the GISAID Initiative, on which this research is based. The information on originating laboratories, submitting laboratories, and authors of SARS-CoV-2 sequence data can be found in Supplementary Table S1. We thank Brad Suchoski, Heidi Gurung, and Prasith Baccam from IEM, Inc. in the United States of America for their help converting our R program into the Julia language, which produced an approximate 50x speedup of our computations. This work was supported by the Japan Agency for Medical Research and Development (grant numbers JP20fk0108535, JP21wm0125008). K.I. received funding JSPS KAKENHI (21H03490). C.P. was supported by the World-leading Innovative and Smart Education Program (1801) from the Ministry of Education, Culture, Sports, Science, and Technology, Japan. The funders had no role in the study design, data collection and analysis, decision to publish, or preparation of the manuscript.

## Author contributions

K.I. designed the study. C.P. collected data and conducted the analysis. C.P. and K.I. wrote the paper.

## Data availability

Supplementary Table S1 contains metadata of all nucleotide sequences of SARS-CoV-2 viruses collected from England during 1^st^ January 2021 to 31^st^ July 2021 used for the analysis.

## Conflict of interest

We declare that there is no conflict of interest.

## Notes

### Competing Interest Statement

The authors have declared no competing interest.

