## Supplementary Information for "Predicting the time course of replacements of SARS-CoV-2 variants using relative reproduction numbers"

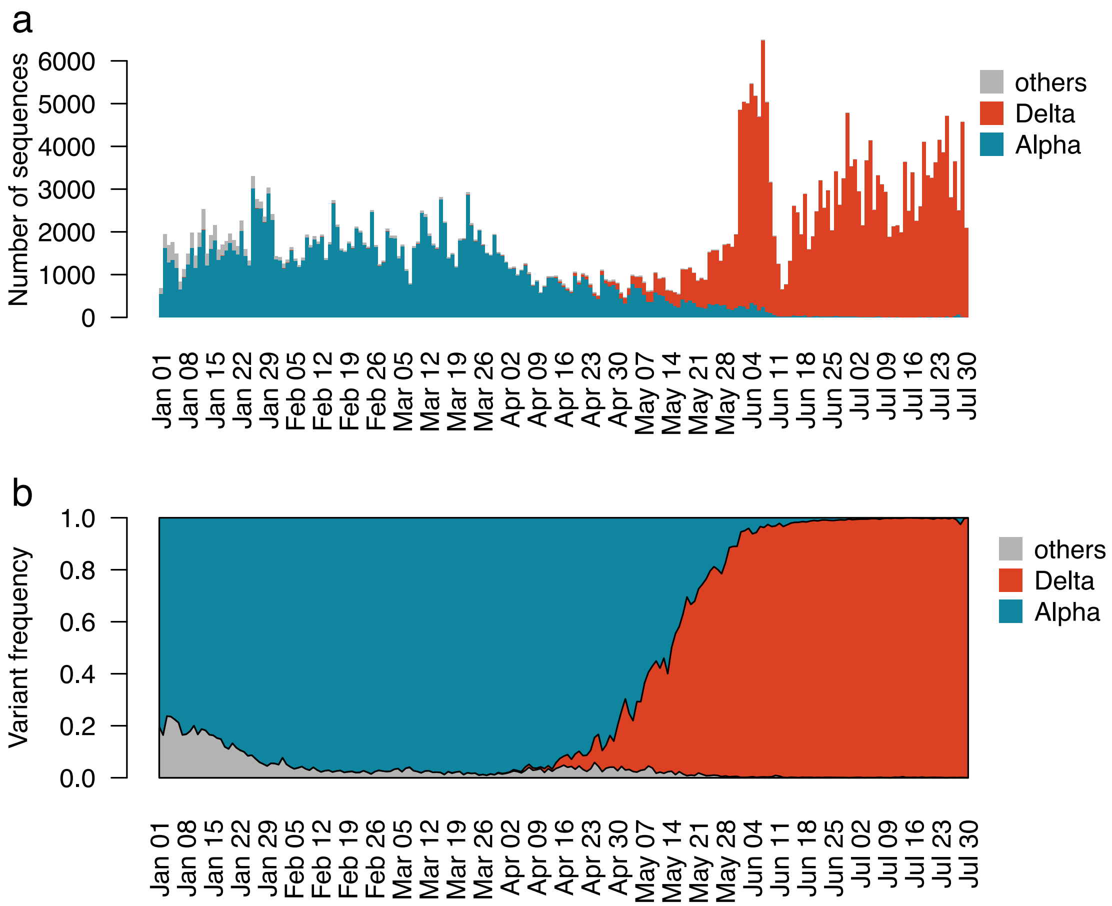


**Supplementary Figure S1. Daily variant frequencies of Alpha (red), Delta (blue), and other variants (gray) in England during 1st January 2021 to 31st July 2021 calculated from nucleotide sequences on the GISAID database**

**Supplementary Table S1. Metadata of nucleotide sequences of SARS-CoV-2 viruses collected from England during 1st January 2021 to 31st July 2021**

**Supplementary Table S2. Parameters estimated by the binomial distribution model and comparison of AIC values with those of the beta-binomial distribution model**

| Observed frequency | Binomial distribution model | | | | Beta-binomial distribution model† | |
|---|---|---|---|---|---|---|
| | $k$ | $q_0$ | Log Likelihood | AIC | AIC | ΔAIC†† |
| 0.05 | 1.90 (1.81, 2.00) | 0.0001 (0.0001, 0.0002) | –86.06 | 176.12 | 170.18 | –5.94 |
| 0.10 | 1.79 (1.73, 1.86) | 0.0002 (0.0002, 0.0003) | –114.37 | 232.75 | 211.71 | –21.04 |
| 0.15 | 1.72 (1.67, 1.76) | 0.0003 (0.0002, 0.0004) | –142.40 | 288.81 | 252.25 | –36.56 |
| 0.20 | 1.74 (1.70, 1.78) | 0.0003 (0.0002, 0.0004) | –159.98 | 323.95 | 281.21 | –42.74 |
| 0.25 | 1.70 (1.67, 1.74) | 0.0003 (0.0003, 0.0004) | –177.35 | 358.71 | 303.96 | –54.74 |
| 0.30 | 1.70 (1.67, 1.73) | 0.0004 (0.0003, 0.0005) | –187.19 | 378.38 | 321.75 | –56.63 |
| 0.35 | 1.70 (1.67, 1.73) | 0.0004 (0.0003, 0.0004) | –190.87 | 385.73 | 329.91 | –55.83 |
| 0.40 | 1.71 (1.68, 1.73) | 0.0003 (0.0003, 0.0004) | –198.37 | 400.73 | 346.75 | –53.99 |
| 0.45 | 1.69 (1.66, 1.71) | 0.0004 (0.0003, 0.0005) | –212.06 | 428.11 | 367.21 | –60.91 |
| 0.50 | 1.67 (1.65, 1.69) | 0.0004 (0.0004, 0.0005) | –233.85 | 471.69 | 386.18 | –85.51 |
| 0.55 | 1.66 (1.64, 1.68) | 0.0005 (0.0004, 0.0005) | –242.85 | 489.71 | 404.18 | –85.53 |
| 0.60 | 1.66 (1.64, 1.68) | 0.0005 (0.0004, 0.0005) | –250.45 | 504.90 | 420.47 | –84.44 |
| 0.65 | 1.67 (1.65, 1.69) | 0.0004 (0.0004, 0.0005) | –259.43 | 522.87 | 431.99 | –90.87 |
| 0.70 | 1.67 (1.65, 1.68) | 0.0004 (0.0004, 0.0005) | –268.59 | 541.19 | 450.39 | –90.80 |
| 0.75 | 1.66 (1.65, 1.68) | 0.0004 (0.0004, 0.0005) | –275.65 | 555.31 | 467.43 | –87.88 |
| 0.80 | 1.67 (1.65, 1.68) | 0.0004 (0.0004, 0.0005) | –286.70 | 577.39 | 494.27 | –83.12 |
| 0.85 | 1.65 (1.64, 1.66) | 0.0005 (0.0004, 0.0006) | –316.19 | 636.37 | 534.58 | –101.79 |
| 0.90 | 1.68 (1.66, 1.69) | 0.0004 (0.0004, 0.0005) | –393.77 | 791.53 | 574.03 | –217.51 |
| 0.95 | 1.72 (1.71, 1.73) | 0.0003 (0.0003, 0.0003) | –521.73 | 1047.47 | 662.66 | –384.81 |
| 1.00 | 1.70 (1.69, 1.71) | 0.0003 (0.0003, 0.0004) | –645.17 | 1294.34 | 867.95 | –426.40 |

AIC refers to the Akaike information criterion of the model.

†AIC values of the beta-binomial model were calculated using parameters in Table 3.

††ΔAIC refers to the difference between AIC values of the beta-binomial distribution and binomial distribution models using the same observation dataset.

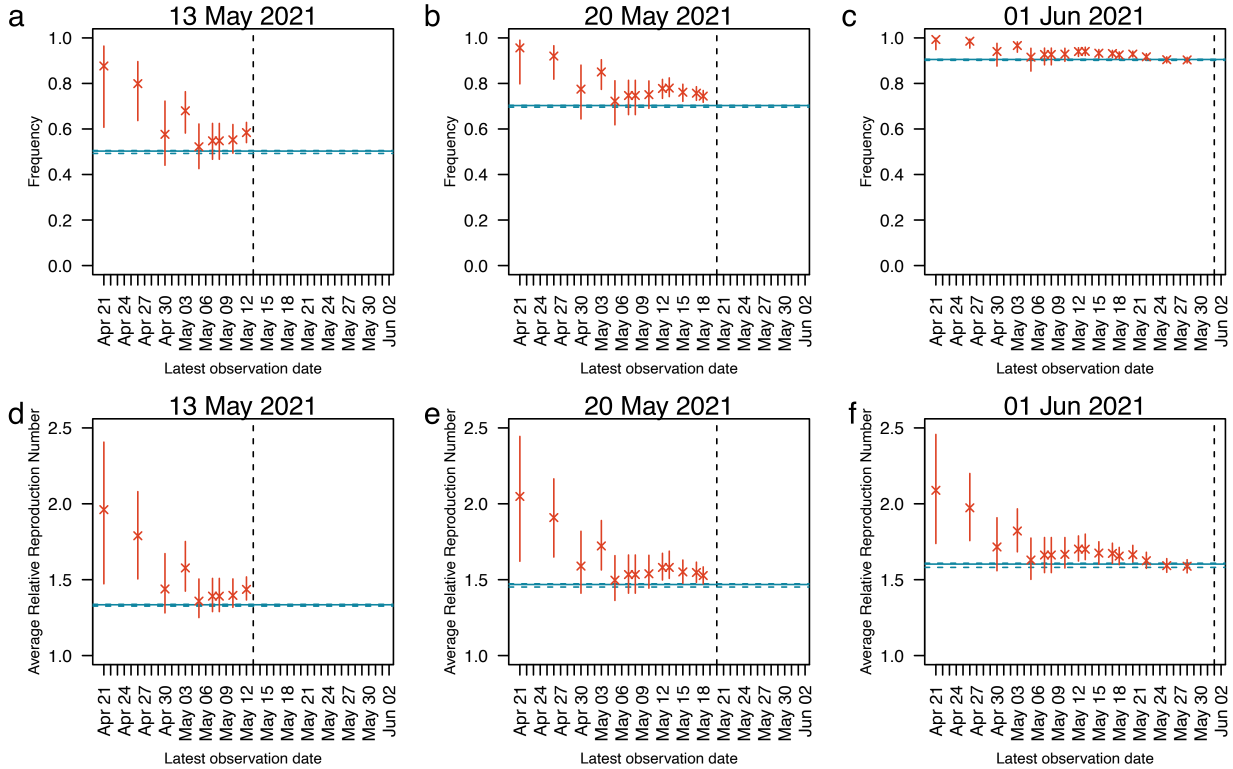


**Supplementary Figure S2. Predictions of population average of relative reproduction number with respect to Alpha with consideration of submission delay.** Nucleotide sequences submitted after the last observation date were not included in the analysis. In each panel, x-axis represents dates until which observations were used in the prediction. Y-axis represents the predicted population average of reproduction number for the date marked by vertical dashed lines (13th May in **a**, 20th May in **b**, and 1st June in **c**). Cross marks represent predicted population average of relative reproduction number with respect to Alpha and vertical bars represent their 95% confidence intervals. The horizontal solid and dashed blue lines represent maximum likelihood estimation and 95% confidence intervals estimated using the entire observations, respectively.

**Supplementary Table S3. Errors of predictions made at frequencies greater than or equal to 0.25 with consideration of submission delay**

| Target date | Number of predictions | Errors in predicted frequency | | Errors in predicted average relative reproduction number | |
|---|---|---|---|---|---|
| | | Median | Maximum absolute error | Median | Maximum absolute error |
| 13th May 2021 | 5 | 0.048 | 0.084 | 0.057 | 0.101 |
| 20th May 2021 | 9 | 0.051 | 0.080 | 0.071 | 0.113 |
| 1st June 2021 | 13 | 0.027 | 0.041 | 0.060 | 0.099 |
